# An MRI Atlas of Regional Brain Vulnerability to Metastatic Disease

**DOI:** 10.64898/2026.06.29.26356888

**Authors:** Joseph I. Turner, Ariel Arias, Allen Fu, Eric K. Oermann, Douglas Kondziolka

**Author notes:** **Corresponding Authors:** Joseph I. Turner, 550 First Avenue, New York, NY 10016, US, **Email:**.

## Abstract

**Background and Objectives:** Are some brain regions intrinsically more vulnerable to metastatic colonization? We sought to characterize the spatial distribution of brain metastases and determine whether regional patterns vary according to primary tumor origin.

**Methods:** We analyzed baseline MRI scans and expert tumor segmentations from 906 patients with 3,492 brain metastases treated with stereotactic radiosurgery. Lesions were normalized to MNI152 standard space and superimposed to generate probabilistic atlases of metastatic occurrence. Regional metastatic burden was quantified using anatomical and vascular atlases. Spatial distributions were additionally compared between lung cancer and melanoma metastases.

**Results:** Metastatic burden was distributed nonuniformly throughout the brain. The cerebellum demonstrated the strongest enrichment relative to its anatomical volume (fold change 1.61, p < 0.001), accompanied by overrepresentation of the vertebrobasilar circulation (fold change 1.49, p < 0.001). Spatial distribution also varied by primary tumor type. Lung cancer metastases demonstrated greater infratentorial involvement than melanoma metastases (16.6% vs. 8.7%, p < 0.05), with a corresponding increase in cerebellar burden (14.8% vs. 6.8%, p < 0.05), whereas melanoma metastases were relatively concentrated within the frontal lobe (37.7% vs. 24.6%, p < 0.01). Infratentorial enrichment was observed across all carcinoma subgroups, with the greatest enrichment seen in gastrointestinal metastases (32.9% infratentorial).

**Conclusion:** Brain metastases exhibit nonrandom spatial distributions, with preferential involvement of posterior and infratentorial structures. Regional patterns vary according to primary tumor origin, supporting the existence of region-specific vulnerability to metastatic disease.

## INTRODUCTION

Brain metastases (BMs) are the most common intracranial tumor^1,2^, with a prevalence approximately ten times greater than that of primary brain malignancies^3^. BMs are observed in 10-20% of adults with systemic cancer^3,4^, often carry a poor prognosis^5^, and confer worse survival than extracranial metastatic disease alone^6^, with outcomes declining further as lesion burden increases^7^. Because cancer cells that successfully colonize the brain undergo distinct genetic and phenotypic adaptations enabling survival within the brain microenvironment, BMs are increasingly recognized as a distinctive CNS disease in their own right, rather than a simple extension of the underlying primary malignancy^8^.

This biological distinctiveness may play a role in determining regional vulnerability to metastatic seeding. Certain primary cancers disproportionately seed the brain relative to others, though the mechanisms driving this organ-level tropism remain incompletely understood^9,10^. Even within the brain itself, metastases are not uniformly distributed, exhibiting regional predilection with a well-documented propensity for posterior and infratentorial structures^11–14^. This pattern has long been recognized clinically in the dictum that a solitary posterior fossa lesion in an adult should be considered a metastasis until proven otherwise^2^.

Characterizing the anatomic distribution of BMs at scale is therefore of considerable interest. Regional predilection patterns encode information about the neurobiology of metastatic seeding, including vascular access, blood-brain barrier vulnerability, and immune surveillance, and may inform mechanistic understanding and therapeutic targeting. Population-level characterization of lesion distribution is also valuable as machine learning-based segmentation models for BMs are developed and validated on large clinical imaging datasets^15^: a probabilistic atlas of lesion occurrence provides distributional priors that can guide model development and help identify anatomical regions where detection performance warrants particular scrutiny. Furthermore, focal neurological dysfunction is among the most common manifestations of brain metastases^2,16–18^, and lesion location is increasingly recognized as an important determinant of clinical outcomes^19–23^. Understanding where brain metastases preferentially occur therefore provides foundation for future studies linking metastatic anatomy to patient outcomes.

Voxel-based lesion mapping (VBLM) provides the methodological foundation for characterizing brain vulnerability to metastatic disease. This method involves normalizing individual patient imaging to a common atlas space and superimposing lesion segmentations across a cohort to reconstruct the probabilistic distribution of lesion occurrence^24^. This approach has been applied to construct spatial atlases for meningiomas^25,26^ and gliomas^27–31^, and, more recently, brain metastases^32^. Building on these prior efforts, we applied VBLM to the largest publicly available brain metastasis imaging dataset^15,33^, comprising 906 patients presenting for first-time stereotactic radiosurgery. We generated cohort-level and etiology-stratified atlases of metastatic distribution, quantified regional metastatic burden, and validated findings using complementary volumetric, patient-level, and lesion-level analyses. Using a public imaging cohort and releasing the resulting atlas maps without restriction, this study provides an open resource for investigating regional vulnerability to brain metastasis.

## METHODS

### Patient Cohort

We analyzed a publicly available dataset of brain metastases^15,33^ (NYUMets), which contains longitudinal clinical and MRI data for patients treated with Gamma Knife stereotactic radiosurgery (SRS). From this dataset, we identified the subset of patients with expert segmentations created for radiosurgery treatment planning, yielding a final cohort of 906 patients with baseline tumor segmentations available for analysis.

### Image Processing and Atlas Construction

Baseline T1-weighted MRI scans and associated tumor segmentations were normalized to the MNI152 standard-space template distributed with FSL^34^ using EasyReg, a deep learning-based deformable registration tool implemented within FreeSurfer^35–37^. Registration quality was verified by flagging abnormally small or large normalized brain volumes and out-of-bounds lesion masks; no registration errors were identified.

Following registration, all tumor segmentations (906 patients; 3,492 lesions) were superimposed in standard space to generate a voxelwise frequency map representing the number of patients with a lesion at each location (**Figure 1**). Normalization by cohort size yielded a probabilistic atlas in which each voxel represented the proportion of patients with a lesion (**Figure 2A,B**). The procedure was repeated for etiology-specific subsets (**Figure 2C**).

**Figure 1:**
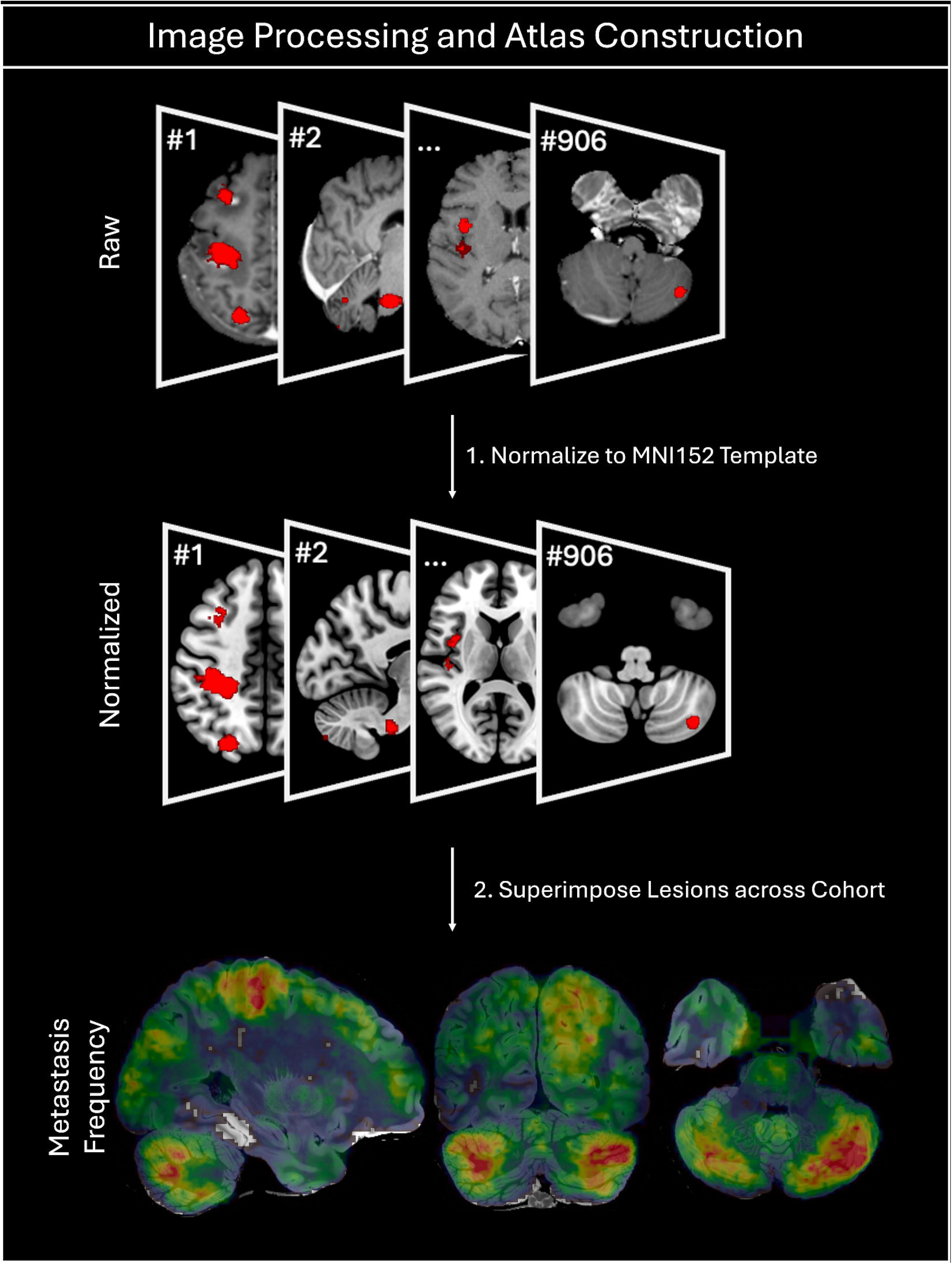
Image Processing and Atlas Construction. Patient scans and lesion segmentations are normalized to the MNI152 atlas, then all lesions are superimposed across the cohort to yield a voxelwise, whole-brain atlas of metastasis frequency. This figure is original to this submission so no credit or license is needed.

**Figure 2:**
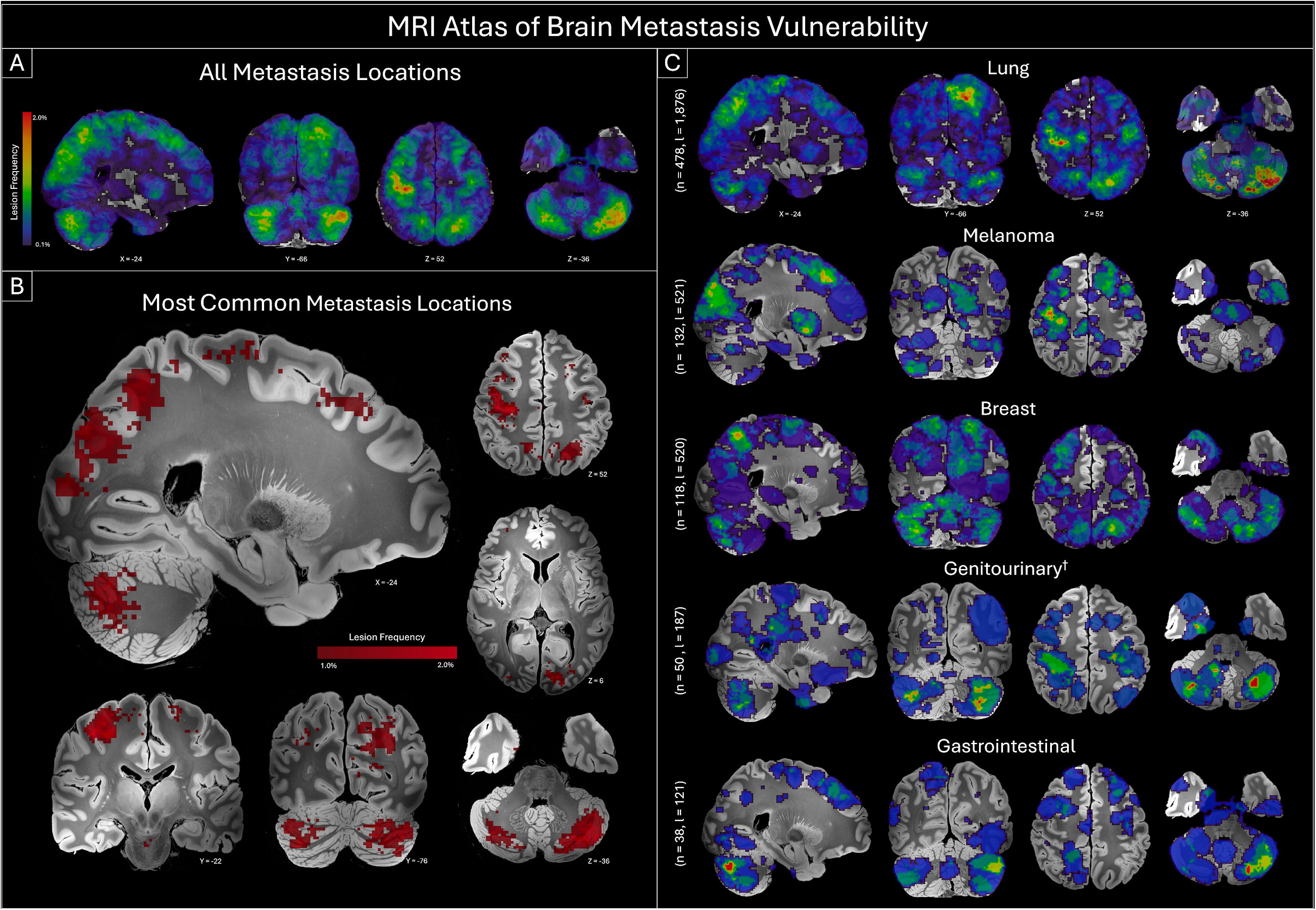
MRI Atlas of Metastasis Burden. **(A)** Unthresholded whole-brain metastasis frequency map. **(B)** Thresholded map displaying high-frequency clusters (affecting >1% of patients), notably at gray-white matter junctions in parietal, occipital, and cerebellar regions. **(C)** Etiology-specific frequency maps for lung, melanoma, breast, and other primary tumors. Coordinates correspond to MNI152 space.This figure is original to this submission so no credit or license is needed. ^†^Genitourinary includes renal, uterine, ovarian, prostate, bladder and testis.

### Regional Lesion Burden Analysis

Tumor segmentations were mapped to standard neuroanatomical regions using the Harvard-Oxford Atlas^38,39^ and a published cerebrovascular territory atlas^40,41^ (**Figure 3A**). Tumors above the cerebellum or brainstem were considered supratentorial. All voxelwise imaging operations were performed in Python using NumPy^42^ and Nilearn^43^.

**Figure 3:**
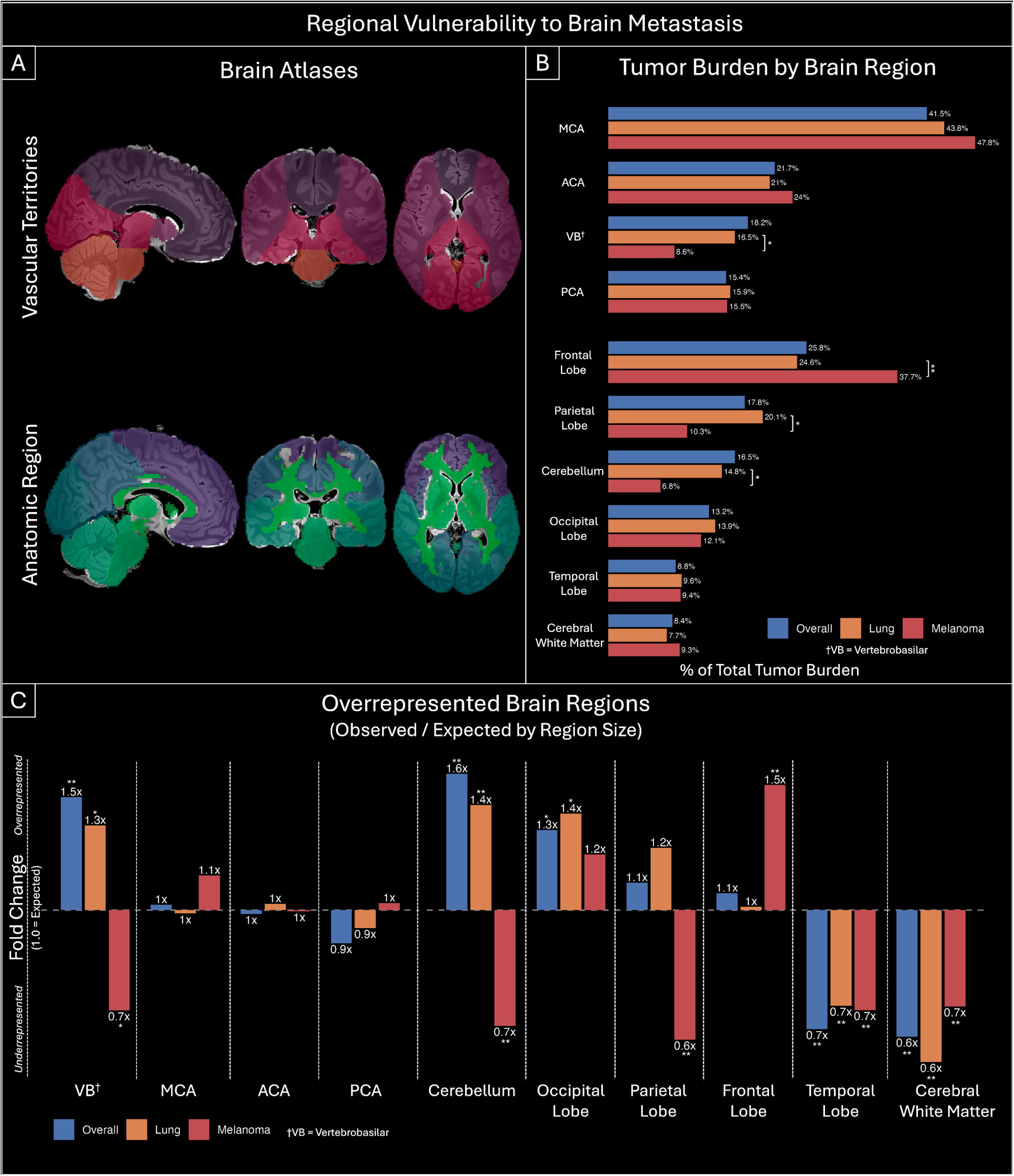
Spatial Distribution by Brain Region. **(A)** Reference atlases defining vascular territories and anatomical lobes. **(B)** Distribution of total cohort metastatic volume across these defined regions. **(C)** Regional tropism, comparing observed versus expected tumor burden. A ratio greater than 1.0 indicates overrepresentation (e.g., vertebrobasilar territory, cerebellum), while less than 1.0 indicates underrepresentation (e.g., temporal lobe). Note melanoma’s distinct preference for the frontal lobe relative to the overall cohort. Significance was determined by comparing the observed fold changes to those derived from a null model of 5000 spatially permuted lesion distributions. This figure is original to this submission so no credit or license is needed.

Our primary measure of regional burden was the cumulative voxelwise tumor volume across the cohort (**Figure 3B**). Aggregating total tumor volume across patients per region avoids the statistical pitfalls of handling boundary-crossing lesions, extreme size variance between individual tumors, and the ambiguity of choosing between patient-level or lesion-level denominators for analyses.

Statistical significance was assessed using two permutation-based approaches (two-sided, 5,000 permutations each). To identify regions with disproportionately high or low tumor burden relative to their physical size **(Figure 3C)**, we generated a null distribution by simulating lesion distributions that preserved an identical number of spatially contiguous lesions with identical volumes as the observed data, but randomizing their intracranial locations (Supplemental Digital Content 1, Methods). Recognizing regional variation in cerebral perfusion, we repeated the analysis of disproportionate tumor burden using a normative cerebral blood flow (CBF) atlas^44^. Rather than comparing tumor volume to regional size, we compared it to a regional perfusion index calculated as the sum of voxelwise CBF values within each region. Significance was assessed using the same permutation framework, except tumor relocation was probability-weighted by normative voxelwise CBF, such that higher-perfusion voxels were proportionally more likely to be selected as relocation sites (**Supplemental Digital Content 2, Figure)**.

To compare spatial distributions between cancer etiologies, we calculated the proportion of total tumor volume attributable to each region and compared observed between-group differences with a null distribution generated by randomly reassigning etiology labels across patients. Uncorrected p values were calculated as the proportion of permutations yielding a result at least as extreme as the observed value. False discovery rate correction (Benjamini-Hochberg) was applied independently across anatomical lobes and vascular territories, and all reported p values are FDR-corrected at α = 0.05. Statistical analyses were performed in Python using SciPy^45^ and statsmodels^46^.

The cumulative volumetric approach was supplemented with patient-level and lesion-level analyses (**Supplemental Digital Content 4, Figure**). At the patient level, we compared the proportion of patients with at least one lesioned voxel in each region using chi-squared tests, and mean per-patient tumor volume per region using Mann-Whitney U tests. At the tumor level, we used chi-squared tests on the proportion of tumor centroids falling within each region.

## RESULTS

### Cohort Characteristics

We analyzed 906 patients with pre-SRS tumor segmentations (**Table 1**). The median patient harbored 2 discrete lesions and 3,488 mm³ of total intracranial tumor volume. Across the cohort, 3,492 individual metastases contributed a cumulative segmented tumor volume of 6.81 × 10 mm³. Lung cancer was the most common primary tumor type (n = 478), followed by melanoma (n = 132), breast cancer (n = 118), and other malignancies (n = 178).

**Table 1.** Baseline Tumor Characteristics. Summary of patient demographics and tumor burden metrics for the entire cohort, stratified by primary etiology and age group. Values represent medians and interquartile ranges (IQR).

|  | N Patients | Median Tumors /Patient | Median Volume/Patient (mm<sup>3</sup>) | Median Volume/Tumor (mm<sup>3</sup>) | Cumulative Tumor Volume (mm<sup>3</sup>) |
| --- | --- | --- | --- | --- | --- |
| **Overall** | 906 | 2 (1, 5) | 3448 (912, 9736) | 488 (192, 1488) | 6,807,344 |
| **Etiology** |  |  |  |  |  |
| Lung | 478 | 2 (1, 5) | 3064 (816, 8120) | 432 (192,1186) | 3,026,968 |
| Melanoma | 132 | 2 (1, 5) | 2484 (486, 8842) | 488 (208, 1424) | 904,360 |
| Breast | 118 | 2.5 (1, 6) | 4900 (1538, 12742) | 464 (184, 1498) | 1,152,872 |
| Genitourinary<sup>†</sup> | 50 | 2 (1, 4) | 5520 (2098, 13994) | 800 (272, 3048) | 623048 |
| Gastrointestinal | 38 | 2 (1, 4) | 6128 (2132.0, 19552) | 1040 (344.0, 4264) | 442240 |
| Other<sup>‡</sup> | 90 | 2 (1, 4) | 3264 (722, 9046) | 560 (208, 2532) | 657856 |
| **Age** |  |  |  |  |  |
| <50 | 147 | 3 (1, 6) | 4644 (1496, 12100) | 492 (200, 1608) | 1,467,064 |
| 50–65 | 339 | 2 (1, 5) | 3160 (916, 9988) | 480 (192,1290) | 2,543,968 |
| 65–80 | 314 | 2 (1, 4) | 3704 (792, 8290) | 468 (192,1450) | 2,040,272 |
| 80+ | 52 | 2.5 (1, 4) | 5212 (1322, 15224) | 656 (192, 1488) | 490,136 |

<sup>†</sup>Genitourinary etiologies included kidney, bladder, prostate, ovarian, uterine, and testis
<sup>‡</sup>Other etiologies included sarcomas, lymphomas, thyroid, and unspecified.

### Atlas Construction

Superimposing all tumor segmentations in standard space generated a whole-cohort probabilistic atlas of metastatic occurrence (**Figure 1, 2A**). High-frequency clusters localized predominantly to cortical gray-white matter junctions throughout the cerebrum and cerebellum, with particularly prominent involvement of parietal, occipital, and cerebellar regions (**Figure 2B**). Etiology-specific atlases were subsequently generated for lung, melanoma, breast, and other primary malignancies, demonstrating distinct spatial distributions across tumor types (**Figure 2C**).

### Regional Vulnerability to Metastatic Disease

To quantify regional susceptibility to metastatic colonization, cumulative voxelwise tumor volume was mapped to published anatomical and vascular atlases (**Figures 3A, 3B**). Regional burden was then compared with a permutation-derived null distribution that accounted for the physical size of each anatomical compartment (**Figure 3C**).

At the tentorial level, the infratentorial compartment contained a significantly greater proportion of metastatic burden than expected by chance (observed 18.3% vs. expected 12.4%, fold change 1.47, p < 0.001). Within this compartment, the cerebellum demonstrated the strongest enrichment across the entire cohort (observed 16.5% vs. expected 10.2%, fold change 1.61, p < 0.001). Consistent with this finding, the vertebrobasilar vascular territory was similarly overrepresented relative to its anatomical volume (observed 18.2% vs. expected 12.3%, fold change 1.49, p < 0.001).

Among supratentorial structures, the occipital lobe exhibited modest enrichment (observed 13.2% vs. expected 9.9%, fold change 1.32, p < 0.05), whereas the temporal lobe (fold change 0.66, p < 0.001) and cerebral white matter (fold change 0.64, p < 0.001) were significantly underrepresented. Together, these findings demonstrate that metastatic burden is distributed nonuniformly throughout the brain, with preferential involvement of posterior and infratentorial regions. Results were highly similar when enrichment was calculated relative to regional cerebral blood flow in place of regional volume, with the same regions remaining significantly over- and underrepresented (**Supplemental Digital Content 2, Figure**).

### Etiology-Specific Patterns of Regional Vulnerability

Lung cancer and melanoma were selected for direct comparison because they represented the two most common etiologies in the cohort and have previously been suggested to exhibit distinct patterns of brain colonization^12,13,32^. The two groups did not differ significantly in median lesion count, median lesion volume, or total patient-level tumor volume. Despite these similarities, their spatial distributions differed substantially (**Figure 3B, 3C**).

At the tentorial level, lung cancer metastases deposited a significantly greater proportion of total tumor volume within the infratentorial compartment than melanoma metastases (16.6% vs. 8.7%, p < 0.05). This difference was driven primarily by the cerebellum, which accounted for 14.8% of total lung metastatic volume compared with 6.8% of melanoma volume (lung fold change 1.44 vs. melanoma fold change 0.67, p < 0.05). A parallel pattern was observed across vascular territories, with lung metastases demonstrating greater involvement of the vertebrobasilar circulation than melanoma (16.5% vs. 8.6%, p < 0.05).

Within supratentorial regions, melanoma metastases were relatively concentrated within the frontal lobe compared with lung cancer (37.7% vs. 24.6%, melanoma fold change 1.55 vs. lung fold change 1.01, p < 0.01). Conversely, lung metastases contributed a larger proportion of tumor volume within the parietal lobe (20.1% vs. 10.3%, p < 0.05).

To assess whether this infratentorial predilection extends beyond lung cancer, we examined additional carcinoma subgroups descriptively. Gastrointestinal malignancies showed the greatest infratentorial enrichment (32.9% vs. 12.4% expected), with breast and genitourinary carcinomas also demonstrating infratentorial overrepresentation (**Supplemental Digital Content 3, Figure**). Given the smaller sample sizes, these findings should be interpreted cautiously but suggest that infratentorial predilection may be a broader feature of carcinomatous metastasis.

### Concordance Across Volumetric, Patient-Level, and Lesion-Level Analyses

To evaluate whether the observed regional patterns were dependent on the choice of analytical unit, we repeated the analysis using both patient-level and lesion-level approaches (**Supplemental Digital Content 4, Figure**). These complementary analyses yielded findings that were broadly consistent with the primary volumetric results. At the patient level, individuals with lung cancer were more likely than those with melanoma to harbor at least one infratentorial metastasis (52.4% vs. 37.6%, χ² = 8.55, p < 0.01) and at least one cerebellar metastasis (49.9% vs. 35.3%, χ² = 8.29, p < 0.05). At the lesion level, lung metastases were more frequently centered within the infratentorial compartment (20.0% vs. 12.7%, χ² = 14.07, p < 0.001), vertebrobasilar territory (20.0% vs. 12.5%, χ² = 15.05, p < 0.001), and cerebellum (18.8% vs. 9.6%, χ² = 23.89, p < 0.001). Melanoma metastases were more frequently centered within the frontal lobe (32.2% vs. 27.6%), although this difference was no longer statistically significant after correction for multiple comparisons (p = 0.09). The concordance of voxelwise volumetric, patient-level, and lesion-level analyses suggests that the observed patterns do not reflect a few outliers with large tumors. Rather, enrichment of cerebellar and vertebrobasilar regions and divergent lung-melanoma distributions reproduced across all analytical frameworks.

## DISCUSSION

In this study, we constructed an atlas of brain metastasis distribution from 906 patients and 3,492 lesions. Consistent with prior reports, metastatic burden was not uniformly distributed throughout the brain, with relative overrepresentation of the cerebellum, occipital lobe, and vertebrobasilar circulation^12,13,32,47,48^ and sparing of cerebral white matter. We further observed that these patterns varied according to primary tumor etiology, with lung cancer metastases preferentially localizing to infratentorial and posterior circulation territories, whereas melanoma metastases demonstrated relative enrichment within frontal regions and avoidance of the cerebellum.

The spatial patterns observed here primarily reflect hematogenous colonization of the brain parenchyma^49^. Mechanical models have historically emphasized the role of cerebral blood flow and vascular anatomy in determining where circulating tumor cells arrest, providing a plausible explanation for the predilection of metastases for gray-white matter junctions^49–51^. However, vascular delivery alone does not fully explain our findings. Because circulating tumor cells from different primary cancers enter the same cerebral arterial circulation, broadly similar spatial distributions would be expected if vascular anatomy were the dominant determinant of lesion location. Instead, we observed consistent differences between lung cancer and melanoma metastases. These findings are consistent with the “seed and soil” hypothesis^52,53^, suggesting that metastatic colonization reflects an interaction between tumor properties and the host brain microenvironment. Experimental studies increasingly demonstrate that brain colonization involves substantially more than passive vascular trapping. Successful metastatic cells exhibit specialized programs that promote brain tropism^54,55^, adapt to the neural microenvironment^56–58^, interact with the blood-brain barrier^59,60^, and engage resident astrocytes and microglia during metastatic progression^61^.

An intriguing possibility raised by these findings is that regional vulnerability may reflect intrinsic heterogeneity of the healthy brain itself. The brain is increasingly recognized as a biologically heterogeneous organ^62^, exhibiting regional variation in vascular architecture^63–66^, cellular composition^67^, metabolic activity^68^, and gene expression^69–73^. Such differences may create ecological niches that vary in their permissiveness to metastatic growth. Future studies integrating atlas-derived vulnerability maps with molecular, cellular, vascular, transcriptomic, and connectomic datasets may help identify the biological features that confer susceptibility to metastatic colonization.

Beyond their biological implications, probabilistic atlases may serve as useful reference resources for future neuro-oncology studies. Because the present atlas was derived from a large publicly available imaging cohort and will itself be made publicly available, it provides a standardized resource for comparing lesion distributions across future datasets and analytical approaches. Increasing evidence across neurology and neuro-oncology suggests that lesion location is an important determinant of clinical outcomes. Neurocognitive dysfunction affects up to 90% of patients with brain metastases prior to treatment^16,17^, and connectome-informed lesion mapping studies have demonstrated that lesion location can predict the cognitive and neurological deficits that emerge following focal brain injury^19–21^. Seizure risk has likewise been linked to lesion location^22,74^, and recent work in diffuse midline glioma suggests that tumor location may independently influence survival^23^. Although this study did not evaluate clinical outcomes directly, the atlas provides a population-derived map of metastatic vulnerability that may facilitate future investigations examining how regional patterns of metastatic involvement relate to neurocognitive morbidity, seizure burden, functional outcomes, and survival. Spatial priors derived from large cohorts may additionally help contextualize lesion distributions observed in individual datasets and inform automated detection and segmentation algorithms.

### Limitations

Several limitations may affect interpretation of these findings. First, this was a single-center retrospective cohort derived from patients treated with stereotactic radiosurgery and may not fully represent the broader population of patients with brain metastases, particularly those managed with surgery, whole-brain radiotherapy, or supportive care alone. Second, histologic and molecular characterization was limited, precluding assessment of spatial differences among biologically distinct tumor subtypes. Third, the atlas reflects radiographically detectable metastases and therefore the end result of successful metastatic colonization rather than the initial distribution of tumor cell seeding. Consequently, the observed patterns likely reflect regional vulnerability to metastatic outgrowth, which may be shaped by immune clearance, cellular dormancy, and other factors occurring after tumor cell deposition. Fourth, although the observed regional predilections remained robust after accounting for regional cerebral blood flow, this study was not designed to identify the biological mechanisms underlying regional vulnerability, which may involve vascular, microenvironmental, immunologic, or blood-brain barrier-related factors. Finally, spatial normalization of brains containing mass lesions is inherently imperfect and may introduce registration error despite the use of modern deformable registration methods.

## CONCLUSIONS

We present a publicly available MRI atlas of brain metastasis distribution that identifies regions of heightened vulnerability to metastatic disease. Regional vulnerability varies according to primary tumor origin, supporting interactions between tumor-specific biology and the host brain microenvironment.

## Supporting information

Supplemental Digital Content 1

Supplemental Digital Content 2

Supplemental Digital Content 3

Supplemental Digital Content 4

## Data Availability

The source imaging and segmentation data analyzed in this study are publicly available through the NYUMets dataset^15,33^. The probabilistic atlas maps generated in this study will be made publicly available via Zenodo upon publication.

## Author Contributions

JIT conceived and designed the study, performed the analyses, interpreted the data, and drafted the manuscript with feedback from all authors. AA and AF contributed to data analysis and interpretation. EKO and DK contributed to study design, interpretation of the findings, and critical manuscript revision.

## Ethics Statement

The study was based exclusively on publicly available, de-identified data and did not involve direct participant interaction or identifiable human subject information.

## Abbreviations

BM: Brain Metastasis
VBLM: Voxel-based Lesion Mapping
SRS: Stereotactic Radiosurgery
ACA: Anterior Cerebral Artery
MCA: Middle Cerebral Artery
PCA: Posterior Cerebral Artery
VB: Vertebrobasilar Circulation
CBF: Cerebral Blood Flow

## REFERENCES

1. Johnson JD, Young B. Demographics of brain metastasis. Neurosurg Clin N Am. 1996;7(3):337–344. doi:10.1016/s1042-3680(18)30365-6

2. Greenberg M. Metastases to the CNS. In: Greenberg’s Handbook of Neurosurgery. Thieme; 2023. doi:10.1055/b000000751

3. Lin X, DeAngelis LM. Treatment of brain metastases. J Clin Oncol. 2015;33(30):3475–3484. doi:10.1200/JCO.2015.60.9503

4. Nayak L, Lee EQ, Wen PY. Epidemiology of brain metastases. Curr Oncol Rep. 2012;14(1):48–54. doi:10.1007/s11912-011-0203-y

5. Hall WA, Djalilian HR, Nussbaum ES, Cho KH. Long-term survival with metastatic cancer to the brain. Med Oncol. 2000;17(4):279–286. doi:10.1007/BF02782192

6. Parker M, Jiang K, Rincon-Torroella J, et al. Epidemiological trends, prognostic factors, and survival outcomes of synchronous brain metastases from 2015 to 2019: a population-based study. Neurooncol Adv. 2023;5(1):vdad015. doi:10.1093/noajnl/vdad015

7. Sperduto PW, Kased N, Roberge D, et al. Summary report on the graded prognostic assessment: an accurate and facile diagnosis-specific tool to estimate survival for patients with brain metastases. J Clin Oncol. 2012;30(4):419–425. doi:10.1200/JCO.2011.38.0527

8. Fares J, Petrosyan E, Dmello C, Lukas RV, Stupp R, Lesniak MS. Rethinking metastatic brain cancer as a CNS disease. Lancet Oncol. 2025;26(2):e111–e121. doi:10.1016/S1470-2045(24)00430-3

9. Yuzhalin AE, Yu D. Brain metastasis organotropism. Cold Spring Harb Perspect Med. 2020;10(5):a037242. doi:10.1101/cshperspect.a037242

10. Obenauf AC, Massagué J. Surviving at a distance: organ specific metastasis. Trends Cancer. 2015;1(1):76–91. doi:10.1016/j.trecan.2015.07.009

11. Delattre JY, Krol G, Thaler HT, Posner JB. Distribution of brain metastases. Arch Neurol. 1988;45(7):741–744. doi:10.1001/archneur.1988.00520310047016

12. Cardinal T, Pangal D, Strickland BA, et al. Anatomical and topographical variations in the distribution of brain metastases based on primary cancer origin and molecular subtypes: a systematic review. Neurooncol Adv. 2022;4(1):vdab170. doi:10.1093/noajnl/vdab170

13. Schroeder T, Bittrich P, Kuhne JF, et al. Mapping distribution of brain metastases: does the primary tumor matter? J Neurooncol. 2020;147(1):229–235. doi:10.1007/s11060-020-03419-6

14. Akeret K, Staartjes VE, Vasella F, et al. Distinct topographic-anatomical patterns in primary and secondary brain tumors and their therapeutic potential. J Neurooncol. 2020;149(1):73–85. doi:10.1007/s11060-020-03574-w

15. Link KE, Schnurman Z, Liu C, et al. Longitudinal deep neural networks for assessing metastatic brain cancer on a large open benchmark. Nat Commun. 2024;15(1):8170. doi:10.1038/s41467-024-52414-2

16. Tucha O, Smely C, Preier M, Lange KW. Cognitive deficits before treatment among patients with brain tumors. Neurosurgery. 2000;47(2):324–333; discussion 333-4. doi:10.1097/00006123-200008000-00011

17. Meyers CA, Smith JA, Bezjak A, et al. Neurocognitive function and progression in patients with brain metastases treated with whole-brain radiation and motexafin gadolinium: results of a randomized phase III trial. J Clin Oncol. 2004;22(1):157–165. doi:10.1200/JCO.2004.05.128

18. Wolpert F, Lareida A, Terziev R, et al. Risk factors for the development of epilepsy in patients with brain metastases. Neuro Oncol. 2020;22(5):718–728. doi:10.1093/neuonc/noz172

19. Boes AD, Prasad S, Liu H, et al. Network localization of neurological symptoms from focal brain lesions. Brain. 2015;138(Pt 10):3061–3075. doi:10.1093/brain/awv228

20. Fox MD. Mapping symptoms to brain networks with the human connectome. N Engl J Med. 2018;379(23):2237–2245. doi:10.1056/NEJMra1706158

21. Salvalaggio A, De Filippo De Grazia M, Zorzi M, Thiebaut de Schotten M, Corbetta M. Post-stroke deficit prediction from lesion and indirect structural and functional disconnection. Brain. 2020;143(7):2173–2188. doi:10.1093/brain/awaa156

22. Schaper FLWVJ, Nordberg J, Cohen AL, et al. Mapping lesion-related epilepsy to a human brain network. JAMA Neurol. 2023;80(9):891–902. doi:10.1001/jamaneurol.2023.1988

23. Sidpra J, Lind V, Cohen AL, et al. A prognostic human brain network for diffuse midline glioma. Nature. Published online June 10, 2026. doi:10.1038/s41586-026-10631-3

24. Bates E, Wilson SM, Saygin AP, et al. Voxel-based lesion-symptom mapping. Nat Neurosci. 2003;6(5):448–450. doi:10.1038/nn1050

25. Patel RV, Yao S, Aguilar Murillo E, Huang RY, Bi WL. Spatial distribution of meningiomas: A magnetic resonance image atlas. Neurosurgery. 2025;96(4):769–778. doi:10.1227/neu.0000000000003149

26. Hosainey SAM, Bouget D, Reinertsen I, et al. Are there predilection sites for intracranial meningioma? A population-based atlas. Neurosurg Rev. 2022;45(2):1543–1552. doi:10.1007/s10143-021-01652-9

27. Bilello M, Akbari H, Da X, et al. Population-based MRI atlases of spatial distribution are specific to patient and tumor characteristics in glioblastoma. NeuroImage Clin. 2016;12:34–40. doi:10.1016/j.nicl.2016.03.007

28. Roux A, Roca P, Edjlali M, et al. MRI atlas of IDH wild-type supratentorial glioblastoma: Probabilistic maps of phenotype, management, and outcomes. Radiology. 2019;293(3):633–643. doi:10.1148/radiol.2019190491

29. Samuel N, Germann J, Yang A, et al. Data-driven probabilistic mapping of the spatial and molecular landscape of glioma. Brain Commun. 2026;8(1):fcaf459. doi:10.1093/braincomms/fcaf459

30. Bao H, Ren P, Yi L, et al. New insights into glioma frequency maps: From genetic and transcriptomic correlate to survival prediction. Int J Cancer. 2023;152(5):998–1012. doi:10.1002/ijc.34336

31. Ren P, Bao H, Wang S, et al. Multi-scale brain attributes contribute to the distribution of diffuse glioma subtypes. Int J Cancer. 2024;155(9):1670–1683. doi:10.1002/ijc.35068

32. Barrios J, Porter E, Capaldi DPI, et al. Multi-institutional atlas of brain metastases informs spatial modeling for precision imaging and personalized therapy. Nat Commun. 2025;16(1):4536. doi:10.1038/s41467-025-59584-7

33. Registry of Open Data on AWS. NYUMets Brain Dataset. https://registry.opendata.aws/nyumets-brain/

34. Jenkinson M, Beckmann CF, Behrens TEJ, Woolrich MW, Smith SM. FSL. Neuroimage. 2012;62(2):782–790. doi:10.1016/j.neuroimage.2011.09.015

35. Fischl B. FreeSurfer. Neuroimage. 2012;62(2):774–781. doi:10.1016/j.neuroimage.2012.01.021

36. Iglesias JE. A ready-to-use machine learning tool for symmetric multi-modality registration of brain MRI. Sci Rep. 2023;13(1):6657. doi:10.1038/s41598-023-33781-0

37. Hoffmann M, Billot B, Greve DN, Iglesias JE, Fischl B, Dalca AV. SynthMorph: Learning contrast-invariant registration without acquired images. IEEE Trans Med Imaging. 2022;41(3):543–558. doi:10.1109/TMI.2021.3116879

38. Collins DL, Holmes CJ, Peters TM, Evans AC. Automatic 3 D model based neuroanatomical segmentation. Hum Brain Mapp. 1995;3(3):190–208. doi:10.1002/hbm.460030304

39. Desikan RS, Ségonne F, Fischl B, et al. An automated labeling system for subdividing the human cerebral cortex on MRI scans into gyral based regions of interest. Neuroimage. 2006;31(3):968–980. doi:10.1016/j.neuroimage.2006.01.021

40. Schirmer MD, Giese AK, Fotiadis P, et al. Vascular Territory template and atlases in MNI space. Published online 2019. doi:10.5281/ZENODO.3379848

41. Schirmer MD, Giese AK, Fotiadis P, et al. Spatial signature of white matter hyperintensities in stroke patients. Front Neurol. 2019;10(208):208. doi:10.3389/fneur.2019.00208

42. Harris CR, Millman KJ, van der Walt SJ, et al. Array programming with NumPy. Nature. 2020;585(7825):357–362. doi:10.1038/s41586-020-2649-2

43. Nilearn contributors, Chamma A, Frau-Pascual A, et al. Nilearn. Zenodo; 2026. doi:10.5281/ZENODO.8397156

44. Meng Z, Ma Y, Zhang W, et al. Age- and sex-specific cerebral blood flow atlases for healthy brain across the lifespan. Sci Data. 2025;12(1):1169. doi:10.1038/s41597-025-05406-w

45. Virtanen P, Gommers R, Oliphant TE, et al. SciPy 1.0: fundamental algorithms for scientific computing in Python. Nat Methods. 2020;17(3):261–272. doi:10.1038/s41592-019-0686-2

46. Seabold S, Perktold J. Statsmodels: Econometric and statistical modeling with python. In: Proceedings of the Python in Science Conference. SciPy; 2010:92–96. doi:10.25080/majora-92bf1922-011

47. Neman J, Franklin M, Madaj Z, et al. Use of predictive spatial modeling to reveal that primary cancers have distinct central nervous system topography patterns of brain metastasis. J Neurosurg. 2022;136(1):88–96. doi:10.3171/2021.1.JNS203536

48. Wang G, Xu J, Qi Y, Xiu J, Li R, Han M. Distribution of brain metastasis from lung cancer. Cancer Manag Res. 2019;11:9331–9338. doi:10.2147/CMAR.S222920

49. Gavrilovic IT, Posner JB. Brain metastases: epidemiology and pathophysiology. J Neurooncol. 2005;75(1):5–14. doi:10.1007/s11060-004-8093-6

50. Hwang TL, Close TP, Grego JM, Brannon WL, Gonzales F. Predilection of brain metastasis in gray and white matter junction and vascular border zones. Cancer. 1996;77(8):1551–1555. doi:10.1002/(SICI)1097-0142(19960415)77:8%3C1551::AID-CNCR19%3E3.0.CO;2-Z

51. Ewing J. Metastasis. In: Neoplastic Diseases: A Treatise on Tumours,.; 1940:62–74.

52. Paget S. The distribution of secondary growths in cancer of the breast. 1889. Cancer Metastasis Rev. 1989;8(2):98–101. doi:10.1016/s0140-6736(00)49915-0

53. Ramakrishna R, Rostomily R. Seed, soil, and beyond: The basic biology of brain metastasis. Surg Neurol Int. 2013;4(Suppl 4):S256–64. doi:10.4103/2152-7806.111303

54. Hoshino A, Costa-Silva B, Shen TL, et al. Tumour exosome integrins determine organotropic metastasis. Nature. 2015;527(7578):329–335. doi:10.1038/nature15756

55. Klotz R, Yu M. Insights into brain metastasis: Recent advances in circulating tumor cell research. Cancer Rep. 2022;5(4):e1239. doi:10.1002/cnr2.1239

56. Ciminera AK, Jandial R, Termini J. Metabolic advantages and vulnerabilities in brain metastases. Clin Exp Metastasis. 2017;34(6-7):401–410. doi:10.1007/s10585-017-9864-8

57. Onwudiwe K, Burchett AA, Datta M. Mechanical and metabolic interplay in the brain metastatic microenvironment. Front Oncol. 2022;12:932285. doi:10.3389/fonc.2022.932285

58. Tyagi A, Wu SY, Watabe K. Metabolism in the progression and metastasis of brain tumors. Cancer Lett. 2022;539(215713):215713. doi:10.1016/j.canlet.2022.215713

59. Karreman MA, Bauer AT, Solecki G, et al. Active remodeling of capillary endothelium via cancer cell-derived MMP9 promotes metastatic brain colonization. Cancer Res. 2023;83(8):1299–1314. doi:10.1158/0008-5472.CAN-22-3964

60. Wrobel JK, Toborek M. Blood-brain barrier remodeling during brain metastasis formation. Mol Med. 2016;22(1):32–40. doi:10.2119/molmed.2015.00207

61. Lorger M, Felding-Habermann B. Capturing changes in the brain microenvironment during initial steps of breast cancer brain metastasis. Am J Pathol. 2010;176(6):2958–2971. doi:10.2353/ajpath.2010.090838

62. Markello RD, Hansen JY, Liu ZQ, et al. Neuromaps: Structural and functional interpretation of brain maps. Nat Methods. 2022;19(11):1472–1479. doi:10.1038/s41592-022-01625-w

63. Wilhelm I, Nyúl-Tóth Á, Suciu M, Hermenean A, Krizbai IA. Heterogeneity of the blood-brain barrier. Tissue Barriers. 2016;4(1):e1143544. doi:10.1080/21688370.2016.1143544

64. Bernier LP, Brunner C, Cottarelli A, Balbi M. Location matters: Navigating regional heterogeneity of the neurovascular unit. Front Cell Neurosci. 2021;15:696540. doi:10.3389/fncel.2021.696540

65. Gulban OF, Stirnberg R, Tse DHY, et al. Whole-brain meso-vein imaging in living humans using fast 7-T MRI. Sci Adv. 2026;12(2):eaea4540. doi:10.1126/sciadv.aea4540

66. Sargent SM, Bonney SK, Li Y, et al. Endothelial structure contributes to heterogeneity in brain capillary diameter. Vasc Biol. 2023;5(1):e230010. doi:10.1530/VB-23-0010

67. Chen X, Huang Y, Huang L, et al. A brain cell atlas integrating single-cell transcriptomes across human brain regions. Nat Med. 2024;30(9):2679–2691. doi:10.1038/s41591-024-03150-z

68. Vaishnavi SN, Vlassenko AG, Rundle MM, Snyder AZ, Mintun MA, Raichle ME. Regional aerobic glycolysis in the human brain. Proc Natl Acad Sci U S A. 2010;107(41):17757–17762. doi:10.1073/pnas.1010459107

69. Hawrylycz MJ, Lein ES, Guillozet-Bongaarts AL, et al. An anatomically comprehensive atlas of the adult human brain transcriptome. Nature. 2012;489(7416):391–399. doi:10.1038/nature11405

70. Hansen JY, Shafiei G, Markello RD, et al. Mapping neurotransmitter systems to the structural and functional organization of the human neocortex. Nat Neurosci. 2022;25(11):1569–1581. doi:10.1038/s41593-022-01186-3

71. Hansen JY, Markello RD, Vogel JW, Seidlitz J, Bzdok D, Misic B. Mapping gene transcription and neurocognition across human neocortex. Nat Hum Behav. 2021;5(9):1240–1250. doi:10.1038/s41562-021-01082-z

72. PsychENCODE Consortium, Akbarian S, Liu C, et al. The PsychENCODE project. Nat Neurosci. 2015;18(12):1707–1712. doi:10.1038/nn.4156

73. Siletti K, Hodge R, Mossi Albiach A, et al. Transcriptomic diversity of cell types across the adult human brain. Science. 2023;382(6667):eadd7046. doi:10.1126/science.add7046

74. Schaper FLWVJ, Morton-Dutton M, Pacheco-Barrios N, et al. Brain lesions causing parkinsonism versus seizures map to opposite brain networks. Brain Commun. 2024;6(3):fcae196. doi:10.1093/braincomms/fcae196

