## Supplemental Digital Content 1 for "An MRI Atlas of Regional Brain Vulnerability to Metastatic Disease"

**Supplemental Digital Content 1. Methods.**

**Spatial permutation testing:** Statistical significance was assessed using empirically generated null distributions. Because volumetric neuroimaging data exhibit substantial spatial autocorrelation and surface-based permutation approaches such as spherical rotations ("spin tests") are not applicable to volumetric lesion maps, we generated null distributions by simulating metastasis patterns directly within the brain volume. This approach approximates the intuitive goal of random spatial relocation while avoiding invalid lesion placements beyond brain boundaries and the central-brain bias introduced by excluding such placements.

For each permutation, the number of simulated metastases was matched exactly to the observed cohort, and each simulated lesion was assigned the volume of a corresponding observed metastasis. Lesions were generated by selecting a seed voxel within the brain mask (excluding ventricles) and iteratively growing a spatially contiguous lesion through neighboring voxels until the target volume was reached. Growth was restricted to voxels within the brain mask (excluding ventricles) and favored expansion into neighboring voxels proximal to the lesion origin, producing compact, approximately convex lesions. The probability of selecting a voxel as the initial seed, as well as the probability of incorporating neighboring voxels during lesion growth, could be weighted by a voxelwise prior map such that selection probability was proportional to voxel intensity.

Simulated lesions preserved the exact number and volume distribution of observed metastases and closely approximated their morphology, exhibiting similar sphericity (0.76 ± 0.11 vs. 0.75 ± 0.09) and convexity (1.58 ± 0.68 vs. 1.39 ± 0.55). As an additional validation, we generated 1,000 simulated metastasis distributions using the empirical metastasis frequency map as the voxelwise prior probability distribution. Simulated tumor frequency maps were highly similar to the observed frequency map derived from patient data (spatial correlation r = 0.80 ± 0.01), demonstrating that the simulation framework reproduces realistic large-scale patterns of metastatic distribution.

Empirical p-values were calculated as the proportion of simulated statistics equal to or exceeding the observed value.
