## Supplemental Digital Content 2 for "An MRI Atlas of Regional Brain Vulnerability to Metastatic Disease"

**Supplemental Digital Content 2. Figure.** Regional tropism analysis adjusted for regional cerebral blood flow. Repeating the permutation analysis using a normative cerebral blood flow atlas yielded results consistent with the primary analysis, with the same regions remaining significantly over- and underrepresented.

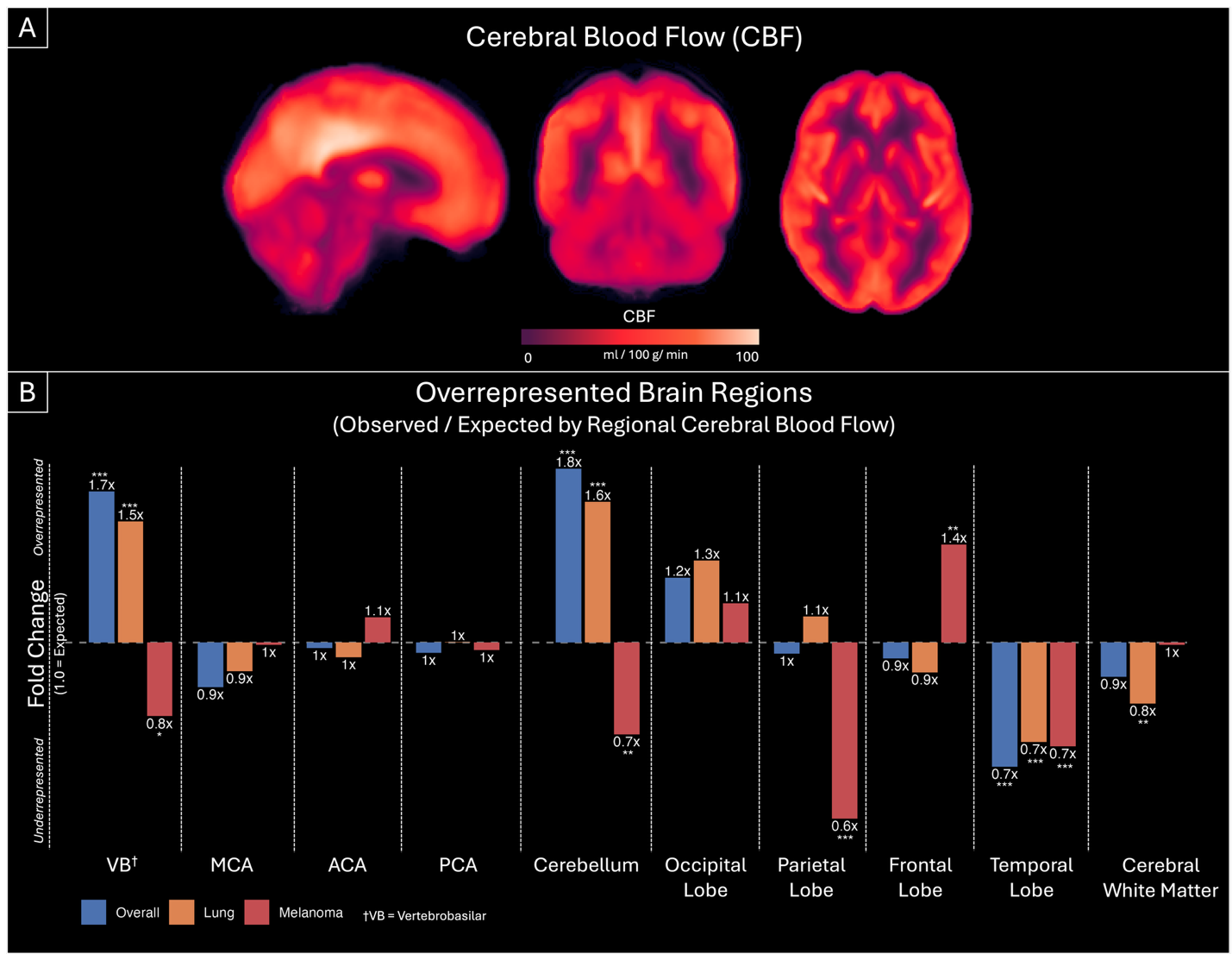
