## Supplemental Digital Content 3 for "An MRI Atlas of Regional Brain Vulnerability to Metastatic Disease"

**Supplemental Digital Content 3. Figure.** Observed versus expected infratentorial metastasis frequency by primary tumor type. Gastrointestinal, breast, genitourinary, and lung carcinomas demonstrated greater-than-expected infratentorial involvement, whereas melanoma did not.

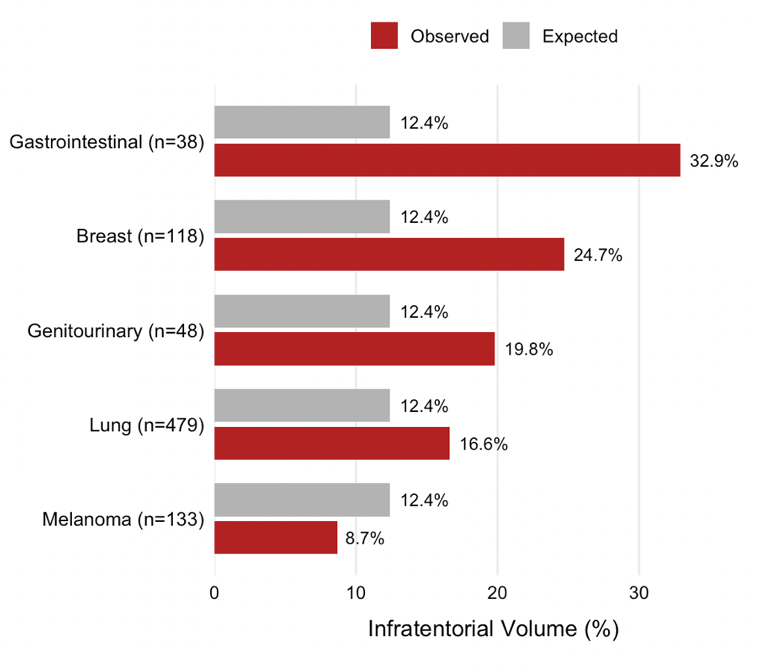
