## Supplemental Digital Content 4 for "An MRI Atlas of Regional Brain Vulnerability to Metastatic Disease"

**Supplemental Digital Content 3. Figure.** Lung vs Melanoma Tumor Locations. **(A)** Patient-level analysis showing the proportion of patients with at least one lesion in each specified compartment, vascular territory, and anatomical lobe. **(B)** Lesion-level analysis showing the proportion of total tumor centroids located within each region. **(C)** Mean tumor mass for lung and melanoma BM patients. Error bars indicate standard deviation. Across metrics, lung metastases demonstrate a significant spatial preference for cerebellar and vertebrobasilar regions relative to melanoma.

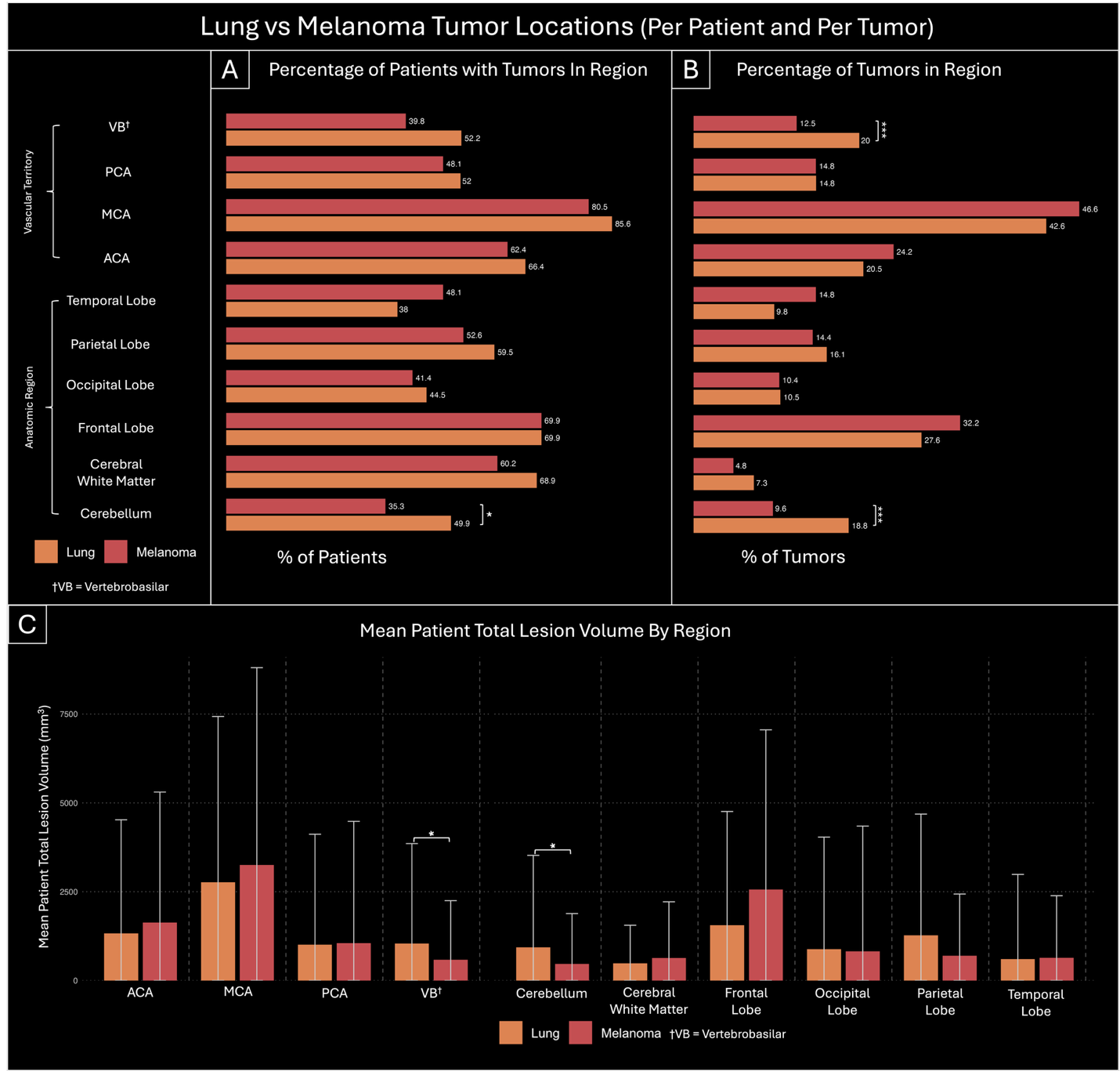
